# Cohort Profile: Health and Attainment of Pupils in a Primary Education National-Cohort (HAPPEN) – A hybrid total population cohort in Wales, UK

**DOI:** 10.1101/2024.05.13.24307272

**Authors:** A Locke, H Jones, M James, E Marchant, J Kennedy, D Burdett, A Bandyopadhyay, G Stratton, S Brophy

**Author notes:** Corresponding Author: Amy Locke.

## Abstract

**Purpose:** HAPPEN is a primary school national cohort which brings together education, health and wellbeing research in line with the Curriculum for Wales framework for health and wellbeing. Health, education and social care data of primary school children are linked and held in the Secure Anonymised Information Linkage (SAIL) Databank. In addition, school-aged children can take part in the HAPPEN Survey throughout the academic year to inform design and implementation of the Health and Wellbeing curriculum area based on their pupils’ needs. There are over 600 schools registered to take part in the HAPPEN Survey. The linkage of health and education records from the HAPPEN national cohort with the HAPPEN Survey responses gives enriched cohort depth and detail which can be used to extrapolate to other schools in Wales. We present the descriptive data available in HAPPEN, and the future expansion plans.

**Participants:** The HAPPEN cohort includes 37,902 primary-aged school children from 2016-July 2023. Of this number, 28,019 can be linked in SAIL with their anonymised linkage field (ALF). In addition, to date (May 2024), HAPPEN Survey data has been captured from over 45,000 children which can in turn be linked to the electronic data. The survey is completed on an ongoing basis and continues to rise by 7000-8000 responses annually.

**Findings to date:** The child cohort is 49% girls, 47% boys (3% prefer not to state their gender and 1% of this data is missing) and 14% are from an ethnic minority background (10% prefer not to state their ethnicity). Initial findings have explored the impact of Covid-19 on wellbeing and play opportunities. As well as a longitudinal exploration of wellbeing throughout the years.

**Future plans:** HAPPEN is an ongoing dynamic cohort of data collection. Access to the cohort is available through SAIL or HDRUK gateway applications. Ongoing research includes the evaluation of interventions for primary school children such as natural experiment methods, non-means tested free school meal roll-out in Wales, interventions to improve physical literacy including changes to the built environment and interventions to improve health and wellbeing of primary school children.

**STRENGTHS AND LIMITATIONS OF THIS STUDY:**

- HAPPEN is a pan-Wales primary school cohort with a specific focus on children’s self-reported health and wellbeing.
- The existence of this cohort and ongoing survey data enables the evaluation of natural experiments
- This cohort facilitates life course research on the health and wellbeing of the children in Wales.
- Missing data or errors in routine and administrative data may be a constraint when discussing population level outcomes for children.
- A potential restriction of HAPPEN is the loss of data pertaining to individuals who relocate outside of Wales during childhood.

## Introduction

Children aged 7-11 typically enjoy good overall health [1]. Nevertheless, there is a concerning prevalence of unhealthy habits among them, including sedentary behaviours, low physical activity, poor dietary intake, and inadequate sleep patterns [2, 3]. Research shows that these unhealthier habits have been increasing in prominence over recent years [4] and these behaviours have significantly intensified since the Covid-19 pandemic [5]. Engaging in these behaviours for extended periods increases the risk of developing negative health outcomes across the lifespan. Recognising the importance of health promotion within educational settings is vital for fostering healthier behaviours and improving educational attainment [6]. However, government policies for school-based health promotion often place this responsibility solely upon schools, resulting in generalised interventions and key health and wellbeing issues being overlooked [7]. Instead, adopting an individualised approach that addresses perceived problems relevant to each school and empowers education settings to adapt to the wants and needs of their own pupils increases the likelihood of successful interventions being embraced [8]. In the context of Wales, the curriculum reform with statutory focus on health and wellbeing offers an array of opportunities for HAPPEN. Also, the curriculum for Wales is an autonomous, school-level curriculum so individualised approaches to health and wellbeing are possible.

The Health and Attainment of Pupils in a Primary Education National Cohort (HAPPEN) is a primary school cohort study which brings together education, health and research in line with Wales’ ongoing education reform and the introduction of the Curriculum for Wales (CfW) from September 2022 [9]. The CfW places statutory focus on health and wellbeing as one of the six Areas of Learning and Experience (AoLE). Furthermore, curriculum design and implementation is done at a school-level, giving schools autonomy to develop a curriculum plan tailored to their pupils’ needs.

The electronic cohort is enhanced by responses to the HAPPEN Survey. The HAPPEN Survey was co-developed following interviews with head teachers who called for a better understanding of school needs in the development of health interventions and advocated for a more holistic approach to improving child health through schools in response to the needs of their own pupils [8, 10-12]. As a result, the HAPPEN Survey was developed alongside staff and pupils to provide an accessible way of capturing a broad range of health and wellbeing behaviours including physical activity, diet and dental health and mental health.

Schools can take part in the HAPPEN Survey throughout the academic year to provide snapshots, track change and evaluate practice. Having completed the survey, schools receive an individual school report aligned with the Curriculum for Wales presenting average data compared to national averages of health and wellbeing in the school. All pupils in year 4, 5 and 6 (ages 7-11) can complete the HAPPEN Survey; an online health and wellbeing questionnaire focused on physical and mental health. Small-scale piloting has been conducted with years 3 (ages 7+). By taking part in the survey, teachers and pupils are empowered to make meaningful changes by gaining a better understanding of pupils’ physical, psychological, emotional and social health. HAPPEN presents the opportunity for pupils to learn and make informed decisions about different aspects of health and wellbeing. The report can be used in school programmes and curriculum design such as Pupil Voice to allow children’s voices to be fully embedded in ways to improve their health.

The Secure Anonymised Information Linkage Databank (SAIL) [13, 14] plays a vital role in facilitating population-based cohorts such as HAPPEN, enabling safe linkage between health and education data. Additional advantages of the HAPPEN dataset and SAIL linkage include the integration of healthcare, social care, and educational datasets to monitor changes over time, support ongoing research, and identify modulating effects on health and educational outcomes.

Previous primary school cohorts have primarily focused on pedagogy and resourcing for schools [15] rather than health, wellbeing, education and wider health linkage with routinely collected data. HAPPEN is the only national primary school network bringing together education, health, child reported information and research. Over 600 schools in the country have registered to take part and over 40,000 pupils have taken part between 2016-2023. Thus, providing an on-going, far reaching primary school cohort. This continued partnership between schools, health professionals and researchers contributes to a more cohesive approach in enhancing child health and well-being across Wales. This document outlines how we have used the HAPPEN Primary School Cohort to build a more comprehensive and complete primary school cohort in Wales. We further set out additional data linkage aspirations using the HAPPEN dataset to continue contributing to children’s health and wellbeing literature.

## Cohort description

The HAPPEN Survey is a self-report health and wellbeing questionnaire focused on physical and mental health completed by pupils in years 4, 5 and 6 (some small piloting work has been done with year 3). The survey asks children about their physical activity and sedentary behaviour (activity levels, motivation active travel, physical literacy, sleep, and concentration), diet and dental health (portions of fruit and veg, fizzy drinks, teeth brushing), mental health (titled “Me and My Feelings” [16] and the Good Childhood Index [17], general wellbeing (autonomy and competence, happiness) and, the local community (happiness with area, walkability to spaces). The survey is conducted in schools whereby all pupils in the year group can take part. This process is governed by opt-out consent. Parents receive the information sheets and opt-out forms prior to the school taking part and can opt their child out of taking part. The school is notified as soon as this happens. HAPPEN has been granted ethical approval by Swansea University’s School of Medicine Research Ethics Board (ref: 7933).

The survey takes between 20-30 minutes to complete independently. Schools can complete the survey at any time during the academic year. Schools can also repeat the survey at different time points should they wish to evaluate practice. Where possible, schools are asked to deliver the survey between Tuesday and Friday, as some questions relate to the previous day and it aims to capture typical school behaviours. It is expected that pupils fill in the survey questions without assistance to make sure they are answering as honestly as possible. If a pupil needs help with reading, a teacher or teaching assistant may read through the question and explain its meaning, but they are asked not to guide the pupil with their answer. HAPPEN has been collecting data since 2016 and has over 45,000 responses to the survey from children across Wales as of May 2024.

## Data-linkage procedures and resources

SAIL employs an Anonymous Linking Field (ALF) to establish connections with other records. Matching of two records arises when their respective ALFs are identical. The matching algorithm was collaboratively designed and assessed by a trustworthy third party and SAIL. The algorithm compares numerous personal identifiers between the received dataset and the Welsh Demographic Service Dataset (WDSD). The linkage is performed based on an National Health Service (NHS) number whenever possible (deterministic linkage). In the absence of an NHS number, a matching algorithm using surname, first name, post code, date of birth and sex is applied against the WDSD (probabilistic linkage). The algorithm’s development ensures a high matching accuracy, with specific thresholds for match accuracy reported to SAIL within the anonymised dataset [8, 9]. A poor matching score indicates incomplete or inaccurate personal information provided by an individual or a lack of registration on the NHS database. The HAPPEN Cohort is updated annually, the most recent upload is available in SAIL from the academic year 2022-2023 (running from September 2022 to July 2023).

## Data sources

Use of HAPPEN data is broad and far reaching. It can be used for longitudinal tracking, cohort studies, evaluations of interventions and natural experiments. With the roll-out of the new Curriculum for Wales and legislation such as the Well-being of Future Generations (Wales) Act 2015 there are a multitude of ways that HAPPEN can contribute to the evaluation and implementation of current practice. The HAPPEN dataset includes linkage to to many datasets including GP and hospital data, national community child health data, education data, ONS births and deaths data, and social care and court data such as Children and Family Court Advisory and Support Service (CAFCASS) family court data.

## Data-linkage hosting environment

Currently, HAPPEN has been linked with core datasets within the SAIL Databank.

The dataset has been used to explore free school meal status as a proxy for deprivation during Covid-19 to predict wellbeing [18]. This was linked in 2021. It has also been linked with the Pathology COVID-19 Daily dataset (PATD) in the same year to explore the association of pre-pandemic (prior to 1 March 2020) health-related behaviours self-reported by children aged 8–11 years during primary school before the COVID-19 pandemic between 1 January 2018 and 28 February 2020, with two outcomes: the odds of ever having a SARS-CoV-2 PCR test and the odds of ever testing positive for SARS-CoV-2 during the period of study [14]. The data has also been linked to the Patient Episode Database for Wales (PEDW). Future research activity has proposed linking to Looked After Children (LACW/LACC), Education (EDUW), Census (CENW), Crown Court Data (CRCO) and the National Community Child Health Database (NCCHD) for collaborative projects with the Ministry of Justice and Health and Care Research Wales. The scope for HAPPEN’s data use in a hosting environment is broad and far-researching.

## Patient and public involvement

HAPPEN was co-developed following interviews with head teachers who called for a better understanding of school needs in the development of health interventions. They also advocated for a more collaborative approach to improving child health through schools. The strong utilisation of consultation, engagement and collaboration has enabled the networks’ success to date. This continued partnership between schools, health professionals and researchers will help provide a more unified approach to improving child health and wellbeing. Patient and public involvement (PPI) is integral to the growth of HAPPEN, underscoring its commitment to inclusivity and participatory research. The inclusion of PPI ensures that members of the public actively participate in shaping the research project.

Valuable input was sought during the design of the surveys from children, parents and school staff. Annual consultations and surveys in schools were conducted about what they found useful. Feedback from this informed iterative development of the survey [19]. The decision-making processes within HAPPEN will prioritise PPI to ensure the inclusion of diverse perspectives. This includes working with local authorities, charities and organisations relating to children’s development, health and wellbeing to ensure a range of priorities are captured. The open text sections of the survey are used to feedback changes children would like to see and feed back to service providers. We undertake focus groups with children on specific topics which are of relevance to them (e.g. free school meals roll out in Wales). To facilitate this approach, HAPPEN follows the Co-production of Research and Strategy (CORDS) standard operating procedure [20] and the UK standards for PPI involvement and National Institute for Health Research (NIHR) guidance from INVOLVE [21] which establishes a framework for meaningful engagement in research.

## Findings to date

### HAPPEN characteristics

The HAPPEN represents a substantial cohort of over 37,902 primary-aged children available in SAIL (of which 28,019 have an ALF; 40% are duplicate entries from children at different time points) and over 45,000 survey responses in school-aged children across Wales as of May 2024. The demographics of the SAIL cohort are in table 1:

**Table 1.** Basic Demographics of HAPPEN Cohort.

| Demographics of HAPPEN Cohort |  |  |  |  |  |
| --- | --- | --- | --- | --- | --- |
| Total Cohort | 37,902 (28,019 with ALF) |  |  |  |  |
| Gender | Boy | Girl | Prefer Not To Disclose | Missing |  |
|  | 47% | 49% | 3% | 1% |  |
| Ethnicity<br>(Since 2019) | White | Black | Asian | Mixed | Prefer Not To Disclose |
|  | 76% | 2% | 4% | 7% | 11% |
| Deprivation<br>(WIMD 2011<br>Quintile) | 1 | 2 | 3 | 4 | 5 |
|  | 23% | 12% | 13% | 10% | 42% |
| Deprivation<br>(WIMD 2019<br>Quintile) | 1 | 2 | 3 | 4 | 5 |
|  | 20% | 17% | 19% | 23% | 21% |
| Year Group | 3 | 4 | 5 | 6 |  |
|  | 2% | 29% | 34% | 35% |  |

The average age of the children is 9.35 years. Recently, the HAPPEN cohort has been used to explore the health and well-being of children during the COVID-19 pandemic. This encompassed a retrospective cohort study employing an online cohort survey conducted between January 2018 to February 2020, in conjunction with routine PCR SARS-CoV-2 test results linked in SAIL. The study explored health-related behaviours in children spanning the period from 2018 to 2020, and their association with being tested and testing positive in 2020 to 2021. Notably, the investigation revealed significant associations between parental health literacy and monitoring behaviours [22]. This research plays a vital role in ensuring the health and wellbeing of young children and school staff and feeds into emerging policy and practice relating to education (for example, conducting rapid research into the impacts of COVID-19 on pupils, teachers and schools, presented to the Welsh Government’s TAG and UK Government’s SAGE groups).

From a pupil’s perspective, free school meal status (FSM) was employed as a proxy for deprivation in an exploratory analysis examining the impact of school closures on the health and well-being. The findings indicated that children eligible for FSM may experience adverse consequences in terms of physical health, including reduced physical activity and suboptimal dietary choices, as a result of prolonged school closures [18].

HAPPEN is also a platform for conducting school-based research, including natural experiment evaluations of school-based programmes. These include outdoor learning [23] and the Daily Mile [24]. Both studies help to provide an evidence base of interventions that can work or be adapted in schools providing recommendations for future implementation and sustainability. It is important that HAPPEN represents the voices of young people in Wales, papers such as the wants and needs of play [25] have helped provide evidence of what children want from play opportunities in their schools namely, more time, space, permission and opportunities for a diverse range of activities. HAPPEN has also been used as a platform to undertake research about teachers and headteachers views [8, 10, 11, 26-28].

## Discussion

### Strengths and limitations

HAPPEN has established a comprehensive cohort which consolidates data from schools, child health and education records. This cohort is supplemented by quantitative and qualitative results from the HAPPEN survey, providing rich insights into details that cannot be obtained through routinely-collected data. The existence of this cohort enables further data linkage, facilitating life course research on the health and well-being of children growing up in Wales. This research aims to generate robust evidence that can influence policy and practice, enabling the implementation of early interventions to promote child health and well-being. This is particularly pertinent in the context of Wales which has undergone significant curriculum reform with the introduction of the Curriculum for Wales. Statutory focus on health and well-being and school-level curriculum design provides opportunities for HAPPEN to inform the design and implementation of schools curriculum tailored to pupils’ health and well-being needs. The cohort exists to empower schools to make meaningful changes by gaining a better understanding of pupil’s physical, psychological, emotional and social health. Without HAPPEN this infrastructure, involving many partners, would not exist. Another strength is that HAPPEN is a platform for conducting wider school-based research including research with teachers, headteachers and allowing intervention evaluations.

One inherent constraint associated with routine and administrative data is the presence of missing data or errors. However, SAIL has established a robust framework that includes a team of skilled analysts and rigorous quality control procedures to address issues such as duplication of patient data entries and minimise instances of missing data. Subsequently, HAPPEN can contribute to the enhancement of clinical reporting practice and bolster the reliability of research findings derived from the cohort. The HAPPEN Survey began as a local project in Swansea, Wales and then expanded regionally and then nationally. The HAPPEN Survey was rolled out across Wales in 2018/2019 and therefore data prior to this point is limited in coverage. Furthermore, a potential constraint of HAPPEN is the loss of data pertaining to individuals who relocate outside of Wales during primary school. It is important to acknowledge that data entry can be subject to human error, and there may still be instances of missing records.

### Research Aspirations

As HAPPEN continues to grow and expand, the network aims to keep its ethos of empowering school and advocating for more holistic working between settings to help improve children’s health and wellbeing. A strong PPI element will be included with this helping to ensure that the work remains aligned with priorities and opportunities that emerge in Wales. The breadth of HAPPEN’s data means there are many opportunities to work collaboratively with external organisations and other academics which is an aspiration for the future. HAPPEN is committed to taking a proactive approach to health and wellbeing helping to create better evidence, opportunities and policy for schools, teachers, parents, organisations and pupils to implement and use.

## Data availability and access

The data for HAPPEN is available in the SAIL Databank at Swansea University, Swansea, UK. All proposals to use SAIL data are subject to review by an independent Information Governance Review Panel (IGRP). This project’s approval code is 0916. Before any data can be accessed, approval must be given by the IGRP. To use HAPPEN data, you need to provide a safe researcher training certificate, a signed data access agreement and IGRP approval.

## Future directions

The HAPPEN cohort enables continuous data collection, facilitating on-going health and wellbeing research including free school meal evaluation, physical literacy and the built environment.

The recent rollout of Universal Free School Meals (UFSM) in Wales [29] and its potential impacts on children’s health outcomes presents a valuable opportunity for investigation within the HAPPEN Cohort. The cohort’s capacity for continuous data collection will facilitate an evaluation of the implementation of UFSM by measuring changes in meal participation, dietary intake (including fruit and vegetable consumption), as well as physical, emotional, and social health via the HAPPEN Survey. The availability of health information via SAIL linkage will also enable us to explore the impacts of UFSM on dietary related illnesses such as respiratory illnesses/infections, gastrointestinal problems, and nutritional deficiencies. This research will provide valuable insights into the potential benefits and implications of UFSM on children’s health and wellbeing outcomes. HAPPEN is a dynamic and evolving cohort which can be aligned to meet research, health, societal and cultural priorities.

## Data availability statement

Data is available upon reasonable request. Researchers can apply for data access by submitting a research application to the SAIL team. The SAIL website provides information on the application process (https://saildatabank.com/data/apply-to-work-with-the-data/). All proposals to use SAIL data are subject to review by an independent Information Governance Review Panel (IGRP). This project’s approval code is 0916. Before any data can be accessed, approval must be given by the IGRP. To use HAPPEN data you need to provide a safe researcher training certificate, a signed data access agreement and IGRP approval.

## Ethics approval

This study involves human participants, HAPPEN has been granted ethical approval by Swansea University’s School of Medicine Ethics Board (ref: 7933). Participants give informed assent to participate in the study before taking part.

## Contributors

All authors provided substantial contributions to the piece of work. AL, HJ, MJ, EM and SB contributed to the conception of this article. AL, HJ, MJ, EM, GS and SB were involved in manuscript writing and revision. MJ EM, and SB were involved in data analysis and interpretation. AL is responsible for the overall content as the guarantor. All authors read and approved the final manuscript. All authors agree to be accountable for all aspects of the work.

